# Pressure-Regulated Ventilator Splitting (PReVentS) – A COVID-19 Response Paradigm from Yale University

**DOI:** 10.1101/2020.04.03.20052217

**Authors:** Micha Sam Brickman Raredon, Clark Fisher, Paul Heerdt, Ranjit Deshpande, Steven Nivison, Elaine Fajardo, Shamsuddin Akhtar, Thomas Raredon, Laura Niklason

**Author notes:** Corresponding author information: Laura E Niklason MD, PhD, Nicholas M. Greene Professor & Vice-Chair, Departments of Anesthesiology & Biomedical Engineering, Yale University.

## Abstract

In the current COVID-19 crisis, the US and many countries in the world are suffering acute shortages of modern ventilators to care for desperately ill patients. Since modern ICU ventilators are powerful devices that can deliver very high gas flow rates and pressures, multiple physicians have attempted to ventilate more than one patient on a single ventilator – so-called “vent splitting”. Early applications of this approach have utilized simple concatenations of ventilator tubing and T-pieces, to provide flow to more than one patient. Additional approaches using custom flow splitters – sometimes made using 3D printing technologies – have also advanced into the clinic with FDA approval. However, heretofore there has been less progress made on controlling individual ventilatory pressures for patients with severe lung disease. Given the inherent variability and instability of lung compliance amongst patients with COVID-19, there remains an important need to provide a means of extending ventilator usefulness to more than one patient, but in a way that provides more tailored pressures that can be titrated over time. In this descriptive report, we provide the basis for a ventilator circuit that can support two patients with individualized peak inspiratory and end-expiratory pressures. The circuit is comprised of exclusively “off the shelf” materials and is inexpensive to produce. The circuit can be used with typical ICU ventilators, and with anesthesia ventilators used in operating rooms. Inspiratory and end-expiratory pressures for each patient can be titrated over time, without changes for one patient affecting the ventilation parameters of the other patient. Using in-line spirometry, individual tidal volumes can be measured for each patient. This Pressure-Regulated Ventilator Splitting (PReVentS) Yale University protocol operates under a pressure-control ventilatory mode, and may function optimally when patients are not triggering breaths from the ventilator.

This method has been tested thus far only in the laboratory with mock lungs, and has not yet been deployed in animals or in patients. However, given the novelty and potential utility of this approach, we deemed it appropriate to provide this information to the broader critical care community at the present time. In coming days and weeks, we will continue to characterize and refine this approach, using large animal models and proof-of-principle human studies.

## The Problem

Recent events had put tremendous stress on our health care system. We have never experienced a healthcare event of such magnitude during our lifetimes. The Centers for Disease Control terms the COVID-19 outbreak a pandemic. Our last pandemic was in 1918, and resulted in nearly 50 million deaths globally and 675,000 deaths in the United States. Events like these put tremendous strain on health care resources. The current pandemic has led to a severe, and growing, shortage of mechanical ventilators.

A survey done by the American Hospital Association in 2009 showed an estimated 62,000 full-featured ventilators. Nearly half of these are in use for pediatric and neonatal patients. Long term vent facilities and vent weaning units will likely have some basic ventilators, though the functionality of these devices is not as complete as typically ICU ventilators. All included, there are approximately 100,000 ventilators in the United States. The Strategic National Stockpile (SNS) had nearly 8,500 ventilators prior to the onset of the COVID-19 epidemic, though now this supply is exhausted. The American Association for Respiratory Care had suggested that the SNS inventory to increase to 11,000–16,000 ventilators in preparation for a severe influenza pandemic. SNS will not suffice for the current pandemic.

The split ventilator technique is not new, and was described first by Drs. Neyman and Babcock in 2006 to ventilate four simulated patients (Neyman 2006). This ventilator ran for 5.5 hours on pressure control, and more than 6 hours on volume control. In 2017, after a mass shooting during a music concert in Nevada, Dr. Kevin Menes used one ventilator to ventilate multiple patients (https://epmonthly.com/article/not-heroes-wear-capes-one-las-vegas-ed-saved-hundreds-lives-worst-mass-shooting-u-s-history/). However, these patients were by and large acute trauma victims and did not manifest the severe respiratory disease that is typically seen with COVID-19 patients.

Recently, a group in Belgium described a pressure control mode for split mechanical ventilation (https://medium.com/@pinsonhannah/a-better-way-of-connecting-multiple-patients-to-a-single-ventilator-fa9cf42679c6) (Pinson 2020). Around the same time, due to the rapid spread of COVID-19 through New York City, Dr. Jeremy Beitler and a team at New York Presbyterian hospital at Columbia University proposed a thorough and thoughtful set of best practices for vent splitting using the Neyman and Babcock method (https://www.gnyha.org/news/working-protocol-for-supporting-two-patients-with-a-single-ventilator/) (Beitler 2020). Beitler and colleagues even began to test this approach on COVID-19 patients (https://www.nytimes.com/2020/03/26/health/coronavirus-ventilator-sharing.html), anticipating that it may soon be necessary to save lives.

However, the approach to vent splitting that has been most widely discussed is ill-suited to the specific challenges of the current COVID-19 pandemic. In the Neyman/Babcock/Beitler approach, multiple patients are attached in parallel to the same ventilator without any intervening valves. Therefore, all patients experience the same airway pressures throughout the respiratory cycle, and ventilator settings cannot be changed for one patient without changing the settings for all others. This approach can be widely used during a mass trauma, where many patients will have healthy lung parenchyma with largely similar ventilation requirements and a straightforward clinical course. COVID-19 patients, on the other hand, can rapidly develop ARDS, and so ventilator requirements can change quickly and in unpredictable ways. Hence, patients who initially are well matched on a single ventilator may no longer be well matched as the day progresses. If the COVID-19 pandemic becomes as widespread as is now feared, affecting hundreds of thousands of US patients, the increase in ICU ventilator production may not be sufficient to keep up with patient demand. This could lead, in turn, to wrenching decisions about which patients should receive ICU ventilator support – heretofore an unthinkable prospect in the modern era.

## A Potential Solution – Pressure Regulated Ventilator Splitting

Here, we propose a circuit design that allows a provider to individualize and independently adjust the inspiratory pressures, tidal volumes, and positive end-expiratory pressures (PEEP) delivered to multiple patients sharing a single ventilator. The method described in this report, the ***P***ressure-***Re***gulated ***Vent***ilator ***S***plitting protocol, or ***PReVentS*** protocol, relies upon currently available and cheaply mass-produced parts that require no special expertise for assembly. The PReVentS ventilator splitting device, and associated guidelines for use, could allow for patient management using the well-established and familiar principles of pressure-controlled ventilation. The PReVentS method could allow for improved care of multiple patients with different disease courses who are forced to share the same ventilator if no better option exists. This method has been tested thus far only in the laboratory with mock lungs, and has not yet been deployed in animals or in patients. However, given the novelty and potential utility of this approach, we deemed it appropriate to provide this information to the broader critical care community at the present time.

The PReVentS protocol has the potential to address all the points brought up by the Multidisciplinary Statement Against Ventilator Splitting, in a statement released on March 26^th^ 2020 (Supplemental Text S1; https://www.sccm.org/Disaster/Joint-Statement-on-Multiple-Patients-Per-Ventilato). To provide a high-level overview of the potential advantages of the PReVentS protocol, we list the specific concerns in the Multidisciplinary Statement with current ventilator splitting approaches, and for each concern we summarize how the PReVentS protocol may ameliorate the problem:

1. *“Volumes would go to the most compliant lung segments”* ***–* In the PReVentS protocol, inspiratory pressures are individualized to the compliance of each patient, and can be adjusted over time**.
2. *“Positive end-expiratory pressure, which is of critical importance in these patients, would be impossible to manage”* **– Positive end-expiratory pressure is individualized to each patient, and can be adjusted over time**.
3. *“Monitoring patients and measuring pulmonary mechanics would be challenging, if not impossible”***-Measuring pulmonary mechanics can be done with a combination of individualized manometry (through either analog manometers or BP transducers) and volume measurement at the ventilator through temporary occlusion of flow to one patient. Additional monitors can provide more detail as available**.
4. *“Alarm monitoring and management would not be feasible”***-Tidal volume alarms on the shared ventilator can be set to detect most types of clinical events, including disconnect, occlusion, inadvertent respiratory effort, and patient compliance changes**.
5. *Individualized management for clinical improvement or deterioration would be impossible”***-Inspiratory and end-expiratory pressures can be individualized and adjusted over time, and there is minimal cross-talk between patients**.
6. *“In the case of cardiac arrest, ventilation to all patients would need to be stopped to allow the change to bag ventilation without aerosolizing the virus and exposing healthcare workers. This circumstance also would alter breath delivery dynamics to the other patients” –* **Laboratory tests simulating cardiac arrest with chest compressions of one patient show retained ventilation of the other patient on the PReVentS circuit. Additionally, removal of one patient from the circuit can be accomplished in a straightforward manner without interruption of the other patient’s ventilation**.
7. *“The added circuit volume defeats the operational self-test (the test fails). The clinician would be required to operate the ventilator without a successful test, adding to errors in the measurement”-* **Individual in-line monitoring of volumes and pressures is the preferred approach when feasible. The self-test can be completed successfully using an equivalent length of respiratory tubing to the PReVentS circuit for appropriate measurement of circuit compliance**.
8. *“Additional external monitoring would be required. The ventilator monitors the average pressures and volumes”* **-Additional monitoring is preferred, and feasible, with the PReVentS approach, including in-line monitoring of tidal volumes and pressures for each patient. Without individual tidal volume monitoring, the ventilator measures the *combined* volumes for both patients, which can provide a sensitive and specific indication of changes in patient and circuit condition**.
9. *“Even if all patients connected to a single ventilator have the same clinical features at initiation, they could deteriorate and recover at different rates, and distribution of gas to each patient would be unequal and unmonitored. The sickest patient would get the smallest tidal volume and the improving patient would get the largest tidal volume”* **– Because the inspiratory and expiratory pressures of each patient are individualized and can be independently adjusted over time, changes in patient condition can be accommodated**.
10. *“the greatest risks occur with the sudden deterioration of a single patient (e*.*g. pneumothorax, kinked endotracheal tube), with the balance of ventilation distributed to other patients” –* **Laboratory tests have shown that, if ventilation to one patient decreases or is cut off, the ventilation to the other patient remains essentially unchanged**.
11. *Finally, there are ethical issues. If the ventilator can be lifesaving for a single individual, using it on more than one patient at a time risks life-threatening treatment failure for all of them” –* **The goal of the PReVentS system is precisely to avoid such ethical issues, by providing high-quality care to more than one patient per ICU ventilator, thereby alleviating critical ventilator shortages**.

More detail on the means by which PReVentS may minimize problems with previous ventilator splitting designs are included in the rest of this document. While the PReVentS approach is not without risks, cannot be safely applied to all patients requiring mechanical ventilation, and should not be applied if better alternatives exist, it could be a valuable addition to the armamentarium of approaches will be needed to minimize mortality if a global pandemic like COVID-19 overwhelms the existing supply of ventilators.

## The PReVentS Ventilator Splitting Circuit – Outline of Approach

Here, we propose a circuit design that allows a health care provider to individualize and independently adjust the inspiratory pressures, PEEPs and tidal volumes delivered to multiple patients who are sharing a single ventilator. This method relies upon currently available and mass-produced parts, that require no special expertise for assembly. The PReVentS ventilator splitting circuit allows for patient management using the well-established and familiar principles of pressure-controlled ventilation. The PReVentS method could allow for improved care of multiple patients having different disease courses, who are forced to share the same ventilator if no better option exists.

In cases of severe lung injury, patients often present with diffuse alveolar injury and high FIO2 requirements. These patients are often ventilated using pressure-regulated ventilation modes, to minimize barotrauma and concomitant exacerbation of lung injury. In such clinical scenarios, patients typically require high levels of PEEP (> 10 cmH2O), moderate to high FIO2, and high inspiratory pressures (> 20 cmH2O). In the most severe cases, patients are heavily sedated and/or paralyzed, with minimal to no spontaneous ventilatory effort, and with all breaths being delivered by controlled ventilation.

The PReVentS protocol is designed for the care of such patients, as a last resort when suitable numbers of contemporary ICU ventilators are not available to ventilate each patient individually. The system described herein is novel in its level of individualized control of both Peak Inspiratory Pressure (PIP) and Positive End-Expiratory Pressure (PEEP), allowing for adaptation to differences in lung compliance and body habitus. Importantly, the system allows for adjustment of PIP and PEEP as support requirements change over time, although respiratory rates will remain the same for each patient. For this setup, is it assumed that patients are sedated and paralyzed to a degree such that they do not trigger breaths delivered from the ventilator. Ventilation triggers can also be set sufficiently high to prevent inadvertent triggering of breaths.

Our proposed approach is not without risks, and cannot be safely applied to *all patients* requiring mechanical ventilation – for example, those patients requiring volume-controlled ventilation are not suitable for the PReVentS approach. Furthermore, ventilator splitting, in general, should not be applied if better alternatives exist. However, we believe that the PReVentS ventilator splitting circuit could provide a valuable addition to the tools needed to minimize mortality if a global pandemic like COVID-19 overwhelms the existing supply of ventilators. This method has been tested thus far only in the laboratory, and has not yet been deployed in animals or in patients.

As of April 2nd, 2020, the PReVentS protocol has been tested extensively in the laboratory using mock lungs (Figure 1). More extensive testing is planned in coming days, including extended application in large animals and brief clinical trials in patients. This description is based upon experimental testing under laboratory conditions only.

**Figure 1:**
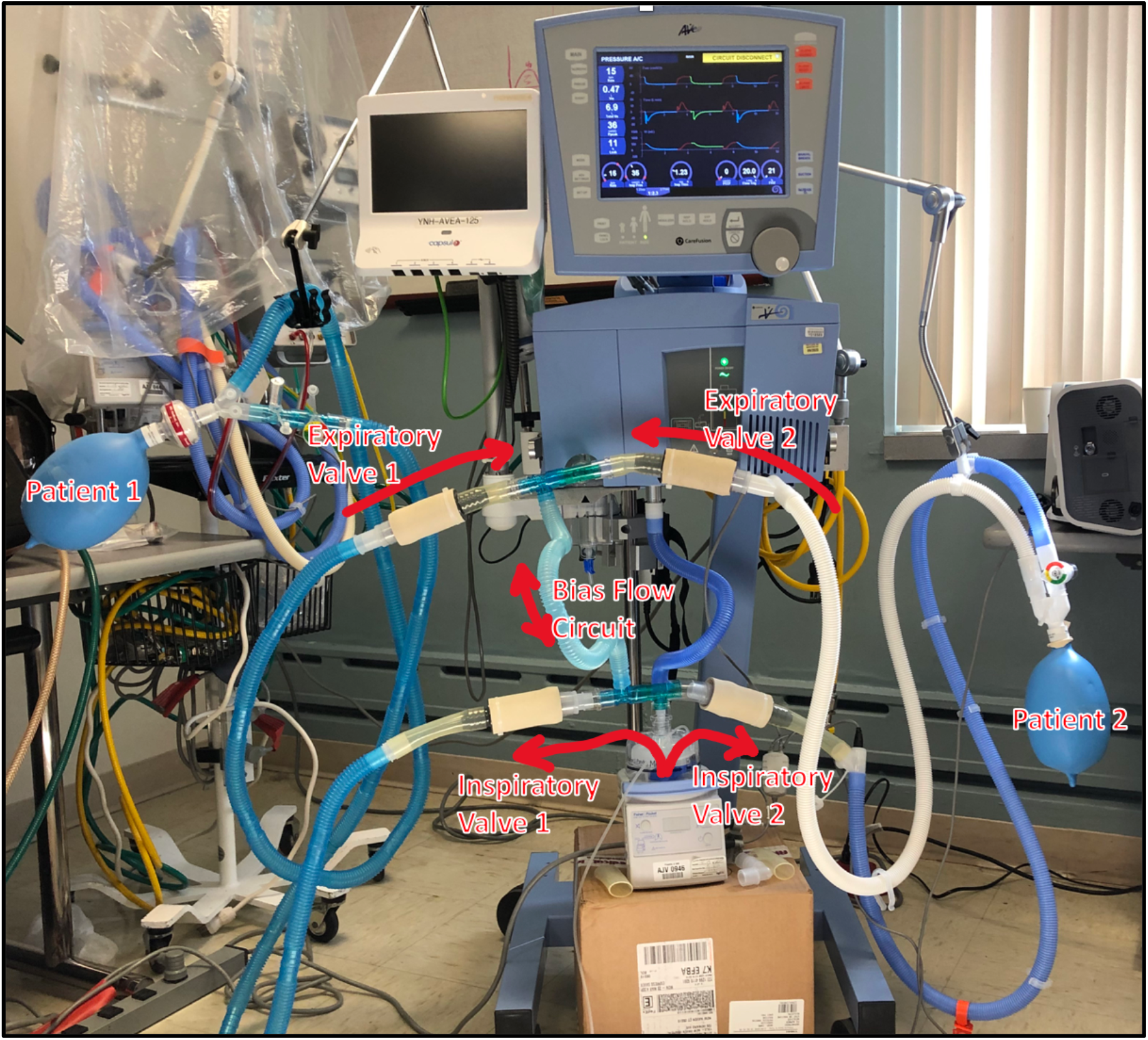
Photograph of the PReVentS Ventilator Splitting Design. Two patients are simulated in this setup. Four pressure-gated inline valves, derived from PEEP valves for bag-valve-mask assemblies, are in the two-patient circuit. Two inspiratory valves control PIP to the two patients. Two expiratory valves control PEEP to the two patients. A “Bias Flow” circuit provides a bypass flow path parallel to the two patient paths to prevent automated ventilator alarming due to high circuit resistance, an artifact which is intrinsic to the placing of in-line pressure-gated valves. **It should be noted that this bypass circumvents a set of alarms which clinicians may be used to relying on during patient care (see below)**.

### Capabilities of the PReVentS Protocol

- Two (or more) patients may be ventilated using a single contemporary ICU ventilator, or using a single contemporary operating room anesthesia machine
- PIP and PEEP are individualized for each patient
- PIP and PEEP can be adjusted for each patient over time, as lung compliance changes
- Valves minimize cross-patient ventilation, even in extreme circumstances such as chest compressions
- Compatible with bacterial and viral filters to minimize infectious cross-contamination
- Compatible with individualized real-time patient spirometry and airway manometry
- Made entirely from off-the-shelf components, no 3D printed or specialty parts needed

### Functional Aspects and Limitations of the PReVentS Protocol

- Pressure-control ventilation mode is optimal and assumed for this design
- Pressure readouts on the ventilator screen reflect unmodified pressures delivered by the ventilator, and are not representative of what each patient is seeing
- PIP and PEEP for each patient are obtained by adding/subtracting each patient’s valve settings from displayed ventilator settings – see below
- Tidal volume readout on the ventilator is total tidal volume for both patients
- Additional monitors may be deployed in-line for each patient, to measure tidal volumes and airway pressures individually and in real time
- FIO2 and respiratory rate are the same for both patients
- Tidal volumes will differ for each patient, depending on PIP, PEEP and lung compliance
- A short circuit from the ventilator outflow to ventilator inflow is necessary to avoid triggering of circuit occlusion alarm and to allow ventilator bias flow
- Because of changes to the expected circuit, ventilator alarms will not always work as expected, and an alternate alarm strategy must be employed (see discussion below)

## Conceptual Basis of the PreVentS Protocol

This protocol uses an ICU ventilator in pressure-controlled mode to create fixed inspiratory and expiratory pressures across parallel patient circuits, and relies upon adjustable valves within each patient circuit to independently modify these driving pressures. This is done using the PEEP valves that are ubiquitous in critical care settings - unidirectional valves that remain open as long as the pressure differential across them exceeds a certain, adjustable threshold. *Because our method uses these valves to adjust inspiratory pressures as well as PEEP, we will hereafter refer to them as pressure-gated valves to minimize confusion*.

Under certain situations, these pressure-gated valves can establish a reliable and adjustable pressure drop from upstream to downstream. If upstream pressure is held fixed, and the pressure differential between upstream and downstream exceeds the valve’s threshold, flow will move through the valve and raise the downstream pressure until the pressure differential no longer exceeds the set threshold. At this point the valve will close, and the pressure downstream will be less than the fixed pressure upstream by *exactly* the threshold pressure of the valve. Similarly, if downstream pressure is held fixed, and the pressure differential exceeds the valve’s threshold, flow will move from upstream to downstream and decrease the upstream pressure until the pressure differential no longer exceeds the set threshold. At this point the valve will close, and the upstream pressure will be greater than the fixed downstream pressure by *exactly* the threshold pressure of the valve. It is by this principle that we have designed the PReVentS system to modulate airway pressures for each patient, by modulating pressure-gated valve settings in the inspiratory and expiratory limbs of the circuit.

Depictions of how the pressure-gated valves are assembled into the PReVentS circuit are displayed below (Figures 1, 2), followed by instructions for circuit assembly and use.

**Figure 2:**
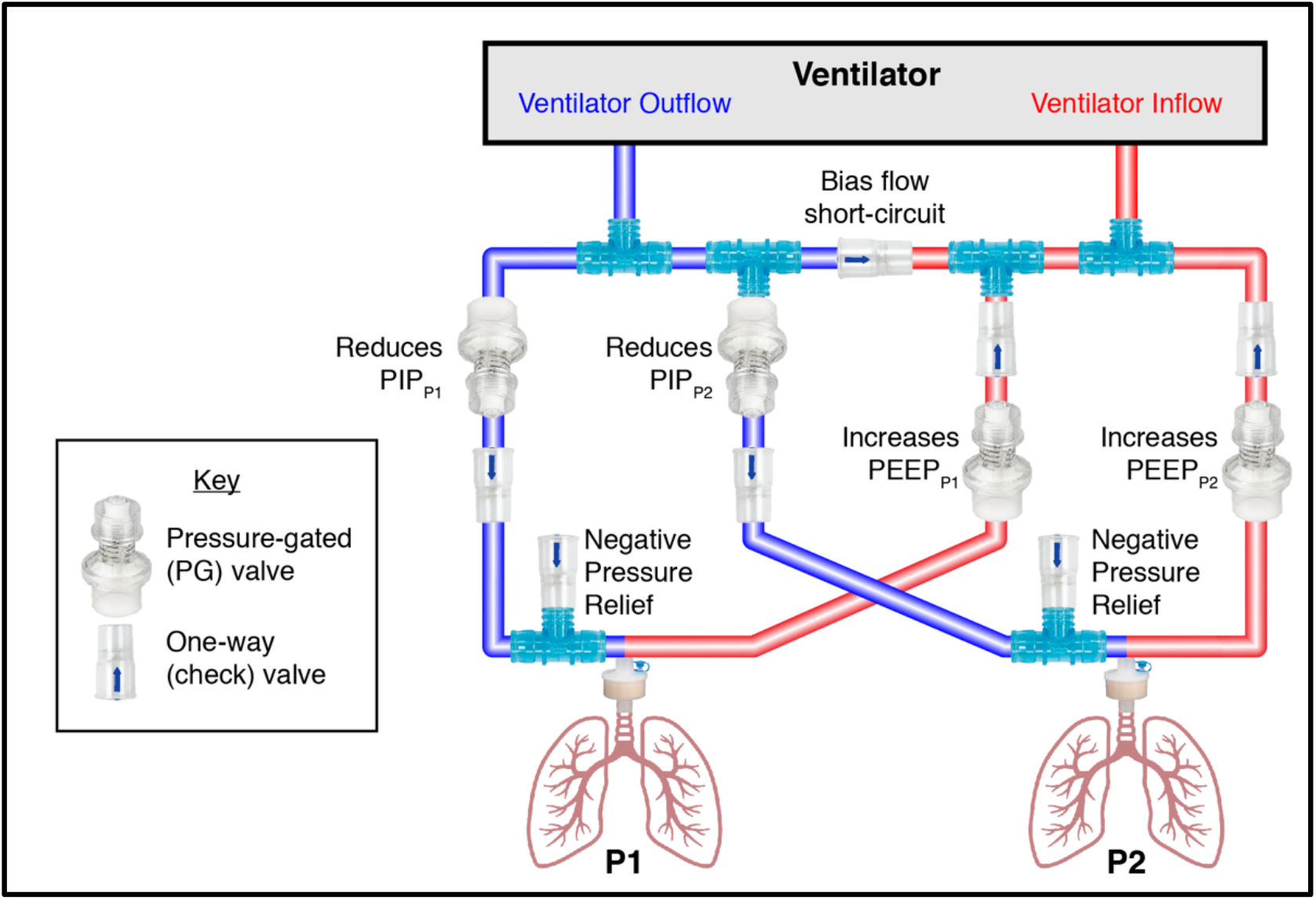
Schematic of PReVentS Ventilator Splitting design. Locations of modified in-line pressure-gated valves, and also one-way check valves to minimize cross-patient ventilation and allow individualized pressure controls, are shown. Functional impact of adjusting each pressure-gated valve is also indicated, for each patient. P1: Patient #1. P2: Patient #2.

### Components (see Figure 2)

– 2 sets of standard ventilator tubing set (Vyaire Medical #1793 or similar)
– 6 standard T-pieces for ventilator tubing (Hudson RCI #1077 or similar)
– 7 in-line check-valves (one-way valve) (Hudson RCI #1664 or similar)
– 2 HME/viral and bacterial filters (ARC Medical Inc. #6000SA or similar)
– 12 ventilator tubing connector cuffs (Hudson RCI 1421 or similar)
– One segment of ventilator tubing for bias flow
– Optional: 2 in-line spirometers (e.g. part # 910004, Medical International Research)
– Parts for modification to create the in-line pressure-gated valve (see Figs 3, 4 below):
  a. 4 disposable PEEP valves, pressure range 1.5 – 20 cmH2O (eg. Ambu Inc., #199003020)
  b. 3/4” inner diameter tubing, Tygon^®^ or other brand
  c. 1” inner diameter tubing, Tygon^®^ or other brand
  d. 1-5/8” inner diameter flexible tubing, or 1-3/4” inner diameter Tygon^®^ tubing
  e. 1-1/2” by 1” tubing reducer

**Figure 3:**
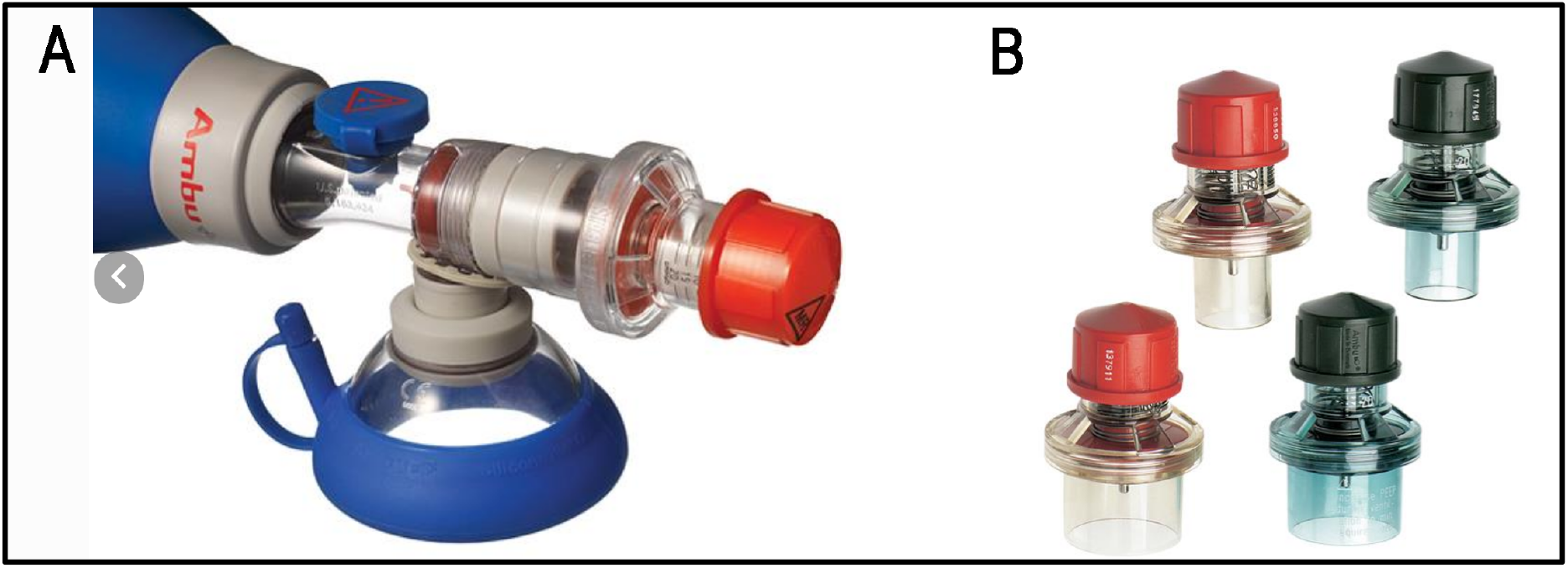
Photographs of single-use Ambu PEEP valves. (A) attached to a bag valve mask assembly; (B) individual PEEP valves that can be used in the PReVentS circuit. (from: https://www.ambuusa.com/products/airway-management/resuscitators/product/peep-valves).

**Figure 4:**
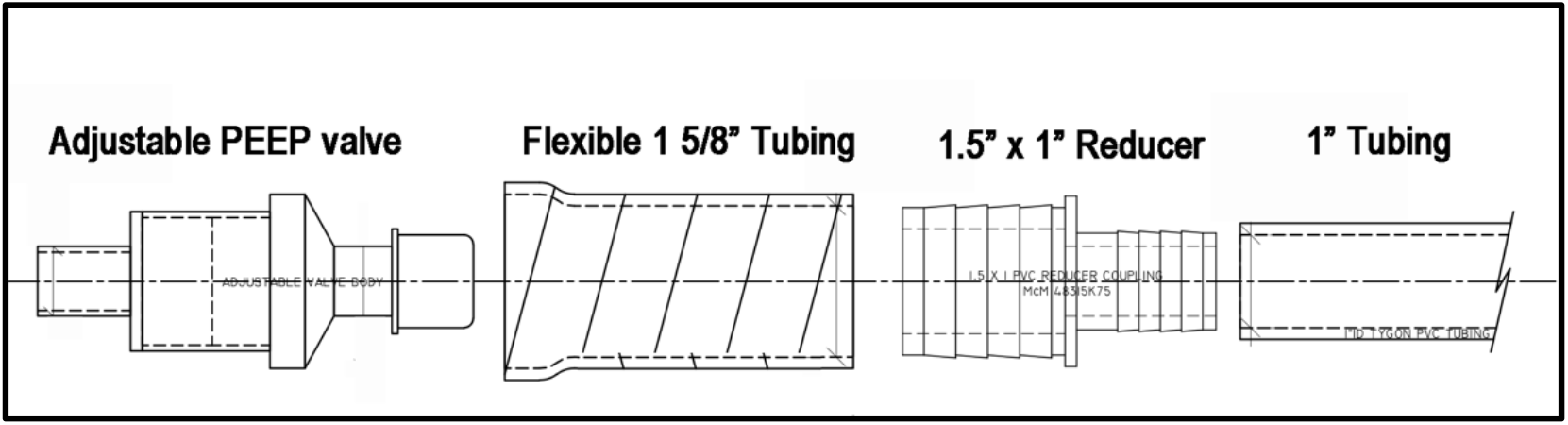
Assembly of In-line PEEP valves. Design drawing of 5 component parts to allow PEEP valves to be positioned in-line in the PReVentS ventilator splitting design. Tubing and Reducer components may be clean, food-grade and durable material, possibly Tygon^®^.

## Instructions for Assembly

Most marketed individual PEEP valves are designed to vent to the outside air, and are not designed to function in-line in the ventilator circuit. Certain in-line PEEP valves do exist on the market, and may be substituted for the design shown here (eg. the BE 142 Magnetic PEEP valve, Instrumentation Industries Inc.). If in-line PEEP valves are not readily available, then existing PEEP valves may be modified to function in-line for this design. Of note, the key to this overall ventilator splitting design is to provide 4 in-line PEEP valves to modulate inspiratory and expiratory pressure for the two patients. The specific type of PEEP valve is not as critical as its functionality.

### I. Step 1: Assembling the In-Line Pressure-Gated Valve

a. Use a disposable PEEP valve (eg. Disposable PEEP valve from Ambu Inc.), that has adjustable PEEP from 1.5 – 20 cmH2O, with a 30mm connector. These valves are typically attached to the outflow of a bag valve mask assembly to apply PEEP to patients undergoing mask ventilation (Figure 3).
b. Cut flexible 1-5/8” ID tubing to 3-3/4” length, and attach to 1-1/2” reducing connector and fasten with ziptie.
c. Cut 1” ID tubing to 2-3/4” length and attach to other end of reducing connector.
d. Set PEEP valve to desired value, and then insert valved end of PEEP valve into the flexible 1-5/8” tubing (see Figure 5 for final valve insertion step). Fasten with ziptie if desired.
e. Repeat these steps 3 more times, to produce a total of 4, in-line pressure-gated valves.
f. Setting of pressure-gated valve values is described below.

**Figure 5:**
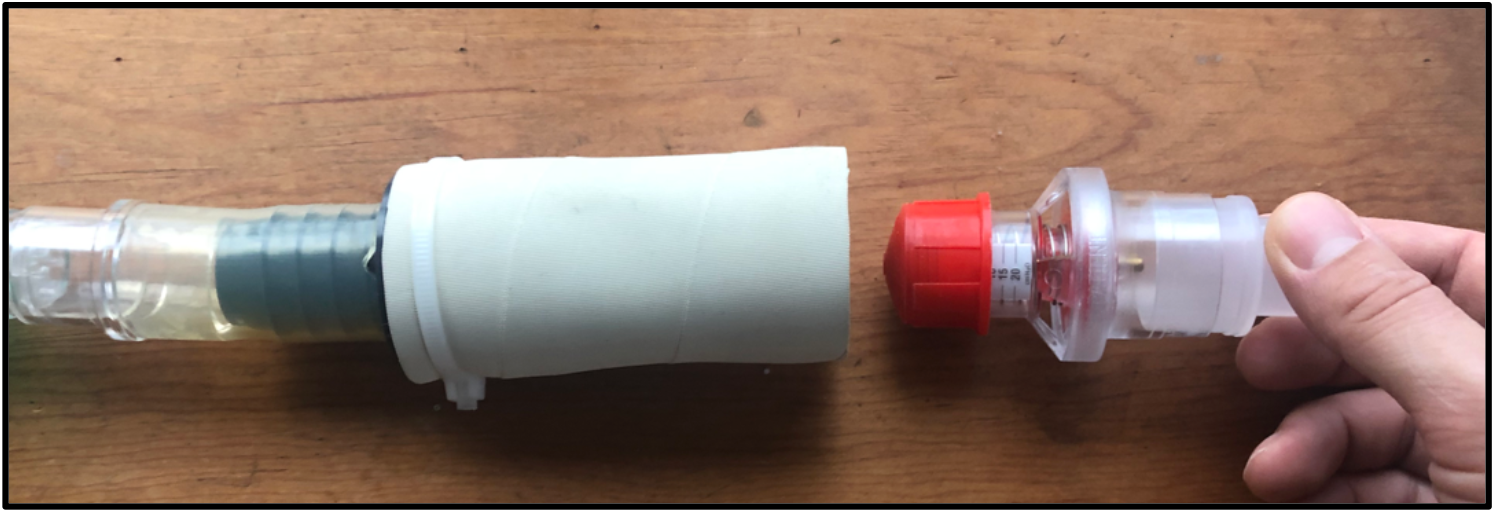
Final step of insertion of PEEP valve into flexible 1-5/8” tubing to produce the in-line PEEP valve for the PReVentS circuit.

### II. Step 2: Connect T-pieces (Figure 6)

a. Attach T pieces to each other into pairs, using 1 connector cuff to attach each pair.
b. Attach connector cuffs to ends of T-pieces as shown in Figure 6.
c. Repeat to create 2 sets of T-pieces.

**Figure 6:**
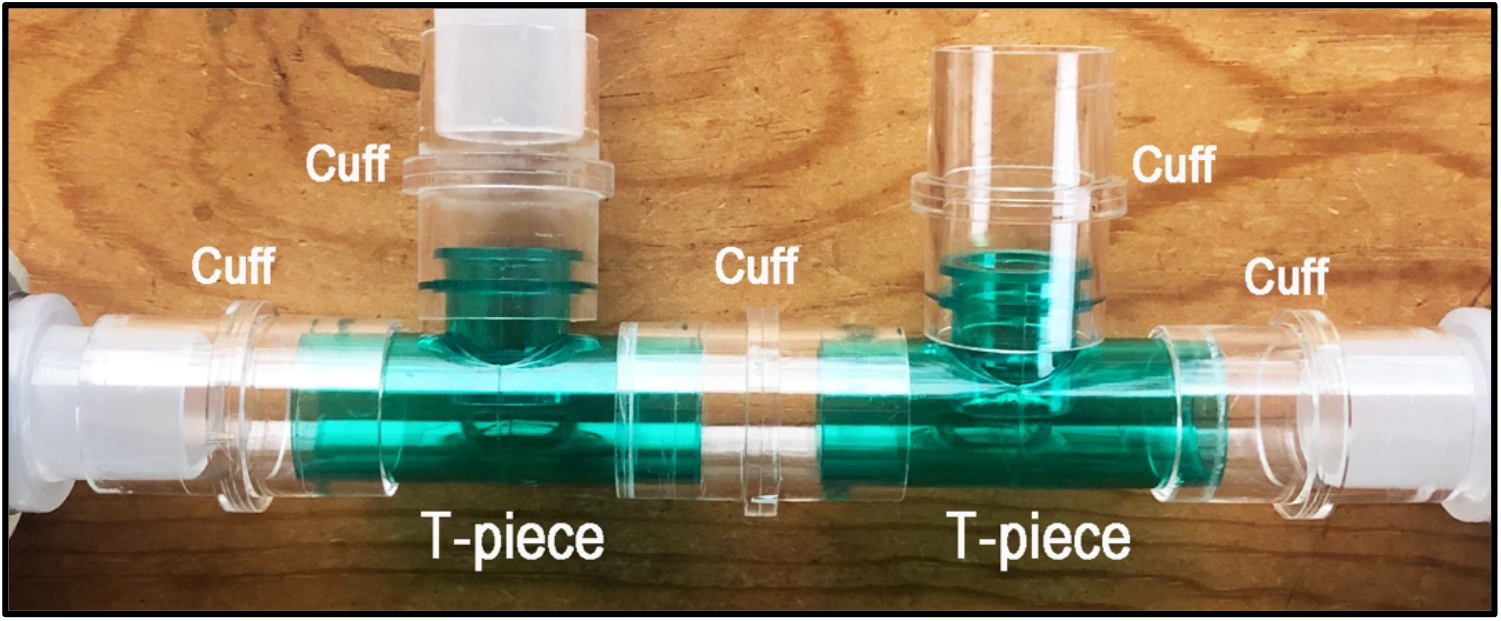
T-piece connections. Two T-pieces are connected using an interposed cuff, and then cuffs are attached to all outlets to provide connections to the circuit.

### III. Step 3: Assemble the Inspiratory Limb of the Circuit (Figure 7)

a. Choose one of the sets of paired T-pieces to be used for the inspiratory limb.
b. Attach an in-line pressure-gated valve, followed by a one-way valve, to each end of the T-assembly. Flow of valves should be *out* of the T-piece pair, in both directions.
c. **TAKE NOTE:** At this point it is important for patient safety to add a negative pressure relief valve into the circuit. While it is not demonstrated in the provided photographs, it is shown in Figure 2 with proper orientation. To do this, add a T-piece after each of the one-way valves from the previous step, and place another one-way valve onto one open port on each of these T-pieces. Flow of all valves must be *inward* toward this T-piece.
d. At the open port on each of these T-pieces, connect the inspiratory limb of a patient circuit.
e. Attach an HME/microbial filter to the end of each patient Y-piece to minimize patient cross-contamination and dissemination of virus into the circuit.

**Figure 7:**
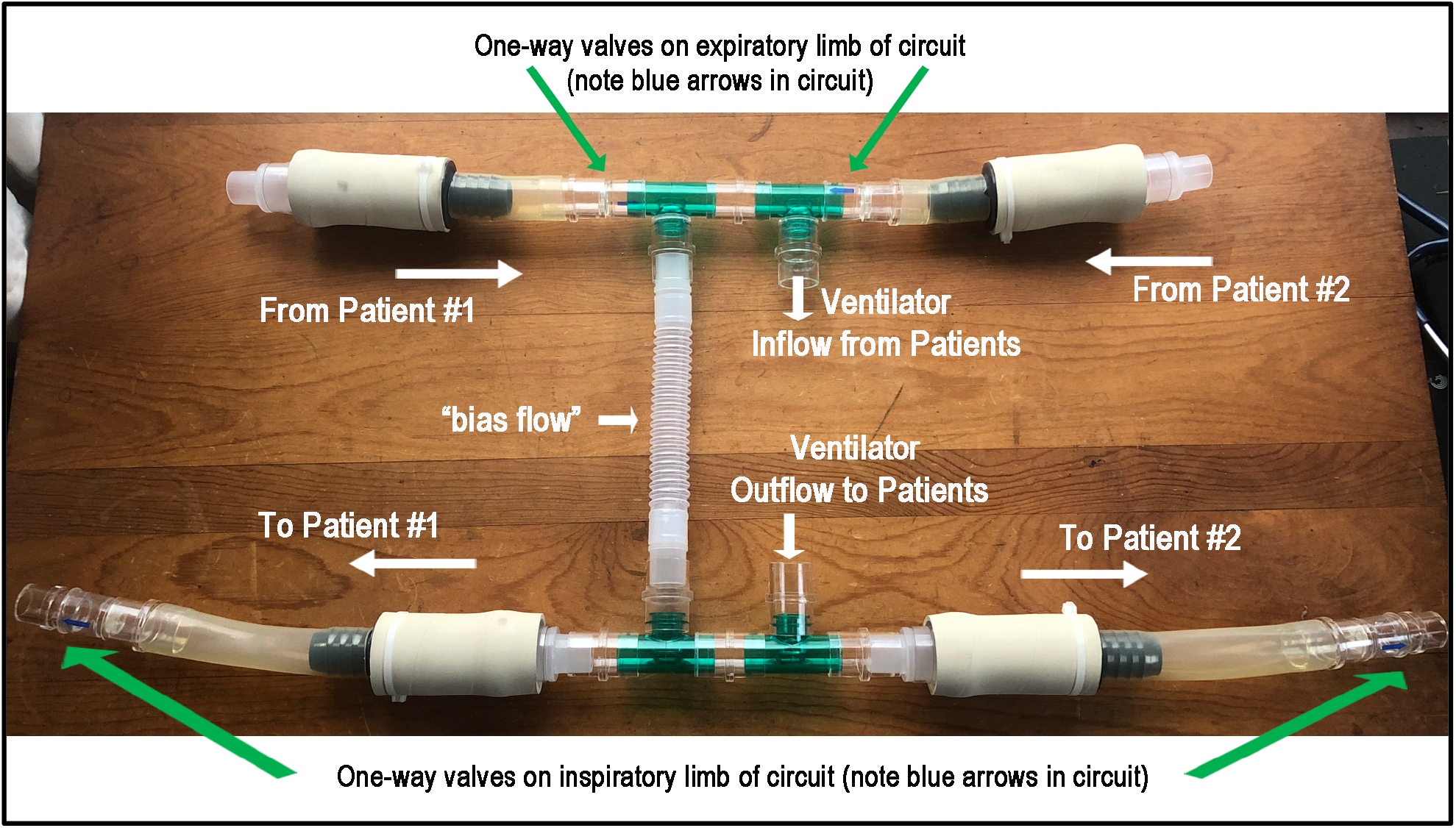
Simplified Overview of the PReVentS Circuit. Flow into and out of the ventilator is shown, next to a “bias flow” circuit which prevents false triggering of the ventilator occlusion alarm. The placement of in-line pressure-gated (PEEP) valves is shown for 2 patients, along with gas flow paths through the in-line valves. Locations of one-way valves are shown in inspiratory and expiratory limbs.

### IV. Step 4: Assemble the Expiratory Limb of the Circuit (Figure 7)

a. Utilize the remaining T-piece assembly
b. Attach a one-way valve followed by a pressure-gated valve to each side of the assembly (flow of valves should point *toward* the T-pieces)
c. Connect expiratory limb of each set of patient tubing to the free end of each of the pressure-gated valves.
d. When available, place in-line flow meters and adapters for pressure transduction between the patient Y-piece and the expiratory limb.

### V. Step 5: Assemble the Inspiratory to expiratory short-circuit (“bias flow”)

Use the short length of respiratory tubing (or an additional connector cuff) to connect the inspiratory T-piece pair to the expiratory T-piece pair. This is necessary on the ICU ventilators in our hospital, so that bias flow is uninterrupted between the inspiratory and expiratory ports of the ventilator, and the ventilator continues to function. See Figure 7. A properly-oriented one-way valve can be added to reduce the chance of rebreathing.

### VI. Step 6: Attach complete circuit to the ventilator

Attach ventilator outflow to the remaining open cuff in the inspiratory limb, and ventilator inflow to the remaining open cuff in the expiratory limb.

## Function of the PReVentS Circuit for Tailoring Ventilatory Pressures

When assembled as described above, the 4 in-line pressure-gated valves combined with 4 in-line one-way valves provide a means of tailoring the ventilatory pressures of two patients, independently. The valve in the inspiratory limb of each patient’s circuit provides a ***step-down*** in Peak Inspiratory Pressure (PIP) equal to its pressure setting. So, the effective PIP for a patient is equal to the PIP set on the ventilator minus the pressure-gated valve setting on the inspiratory limb of the circuit supplying that patient (Figure 8).

**Figure 8:**
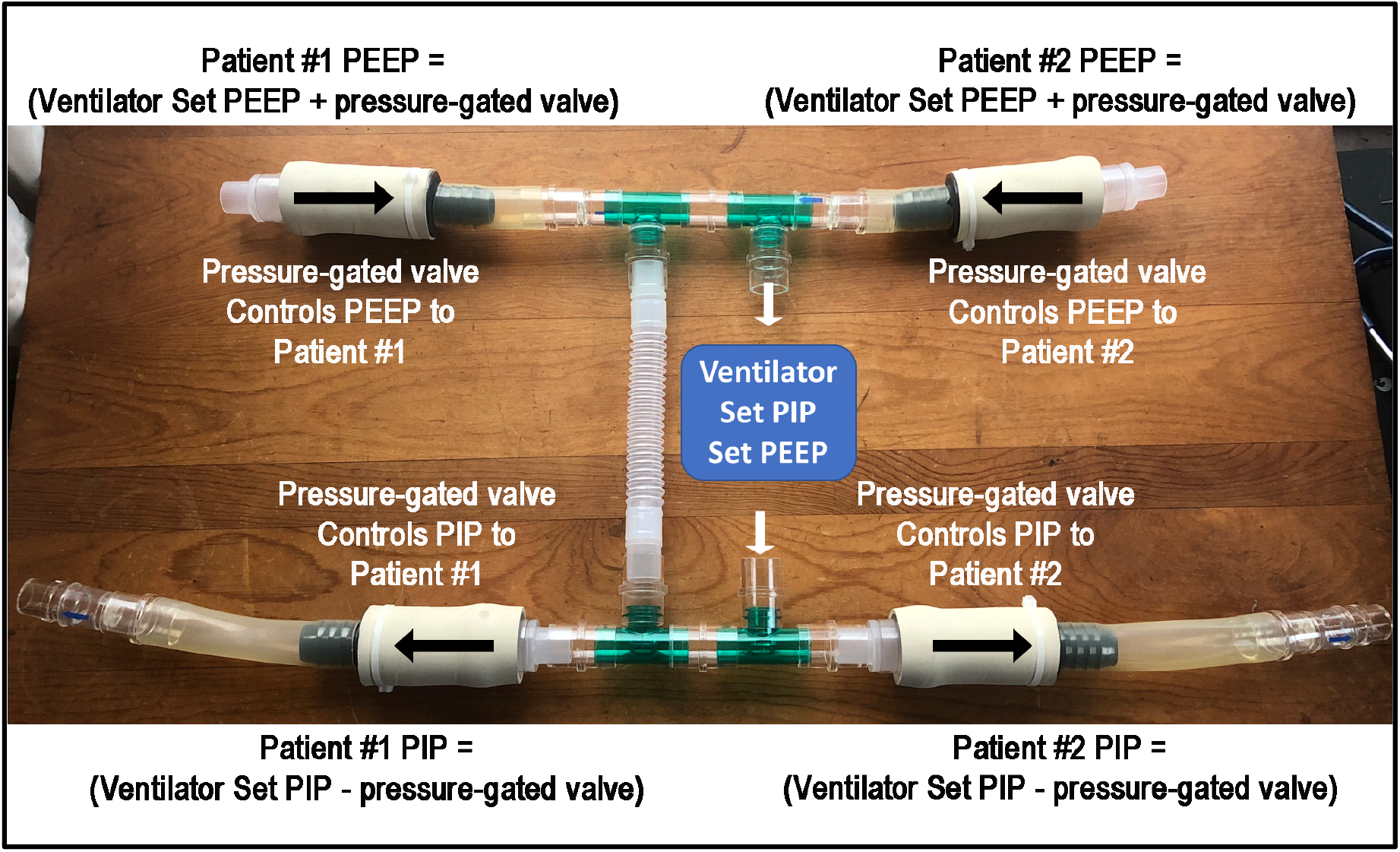
Flow paths of ventilation from the ventilator to each patient. The ventilator provides a single setting of PIP and PEEP to the circuit. The inspiratory limb of the circuit (bottom of figure) contains two in-line pressure-gated valves that control the PIP to patients #1 and #2. The expiratory limb of the circuit (top of figure) contains two in-line pressure-gated valves that control the PEEP to patients #1 and #2.

Conversely, the pressure-gated valve on the expiratory limb of each patient’s circuit provides a ***step-up*** in Positive End-Expiratory Pressure. So, the effective Positive End-Expiratory Pressure for a patient is equal to sum of the PEEP setting on the ventilator, and the setting on the pressure-gated valve in the expiratory limb of that patient’s circuit.

As an example of how the PReVentS ventilator splitting circuit might function for two patients, specific pressures are provided in Figure 9. In this example, Patient #1 has very poor lung compliance and a high PEEP requirement: to maintain adequate minute ventilation and alveolar recruitment, this patient requires a PIP of 35 cmH_2_O and a PEEP of 20 cmH_2_O. In contrast, Patient #2 has less-severe lung disease, requiring less ventilatory support overall, and needs a PIP of only 25 cm H_2_O and a PEEP of 5 cm H_2_O (Figure 9).

**Figure 9:**
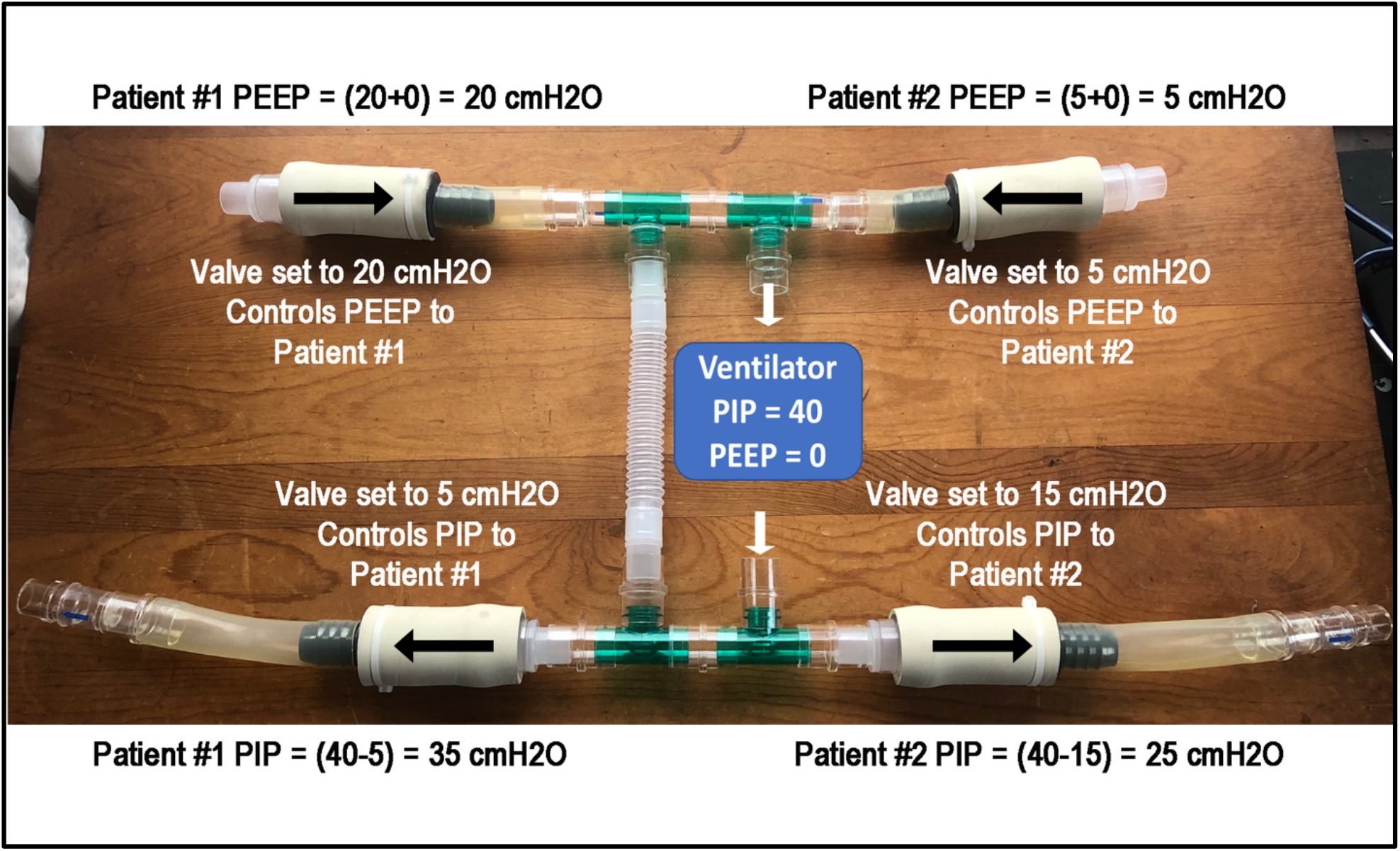
Example of Patient #1 and #2 ventilatory pressures. In this example, the ventilator is set at a PIP of 40 and a PEEP of 0 cmH_2_O. Inspiratory flow at a pressure of 40 cmH_2_O crosses the two in-line PEEP valves near the bottom of the image, resulting in pressure drops of 5 cmH_2_O for Patient #1, and a drop of 15 cmH_2_O for Patient #2. Conversely, on the expiratory limb, the pressure-gated valves provide an effective PEEP of 20 cmH_2_O for Patient #1, and 5 cmH_2_O for Patient #2.

To obtain these target pressures for each patient, the ventilator and PEEP valves would be set up as described below. The ventilator would be set to pressure-controlled mode, with a ventilator PIP of 40 cmH_2_O and a ventilator PEEP of 0 cm H_2_O (Figure 8). Gas from the ventilator would flow into the inspiratory limb of the circuit, where it would flow across two pressure-gated valves. For Patient #1, the inspiratory pressure-gated valve would be set at 5 cmH_2_O, resulting in a pressure drop across the valve of 5 cmH_2_O – therefore, Patient #1 receives a PIP of 35 cmH_2_O. In contrast, for Patient #2 the pressure-gated valve would be set at 15 cmH_2_O, resulting in a drop of 15 cmH_2_O pressure from the ventilator – therefore, Patient #2 receives a PIP of 25 cmH_2_O (Figure 9).

The converse is true on the expiratory limb of the circuit. For Patient #1, the in-line expiratory PEEP valve would provide 15 cmH_2_O of PEEP. For Patient #2, the in-line expiratory PEEP valve would provide 5 cmH_2_O of PEEP. Since the ventilator is set with a PEEP of 0 cmH_2_O, the total PEEP experienced by each Patient is simply the value set for each in-line expiratory pressure-gated valve. However, if the PEEP on the ventilator were set at a number higher than zero – say, at 5 cmH_2_O – then the PEEP experienced by each patient would be the sum of the ventilator set PEEP and the setting of the pressure-gated valve on the expiratory limb of the patient’s circuit.

## Laboratory Studies of the PReVentS Circuit

To document proof-of-principle for the PReVentS circuit, and to test its ability to separately control ventilatory pressures for two patients, we assembled the circuit attached to a standard ICU ventilator. Standard test lungs (eg. BioMed Device # 1020 or similar) were used to simulate patient lungs. To simulate a decrease in lung compliance, we applied rubber bands around one of the test lungs to increase their stiffness. Figures 10-14 below show data obtained from the PReVentS circuit as described above. With the current hand-made valves it is necessary to briefly open the circuit to make valve adjustments. We are working to update the valve design so that pressures can be set without making any disconnects in the PReVentS circuit.

**Figure 10:**
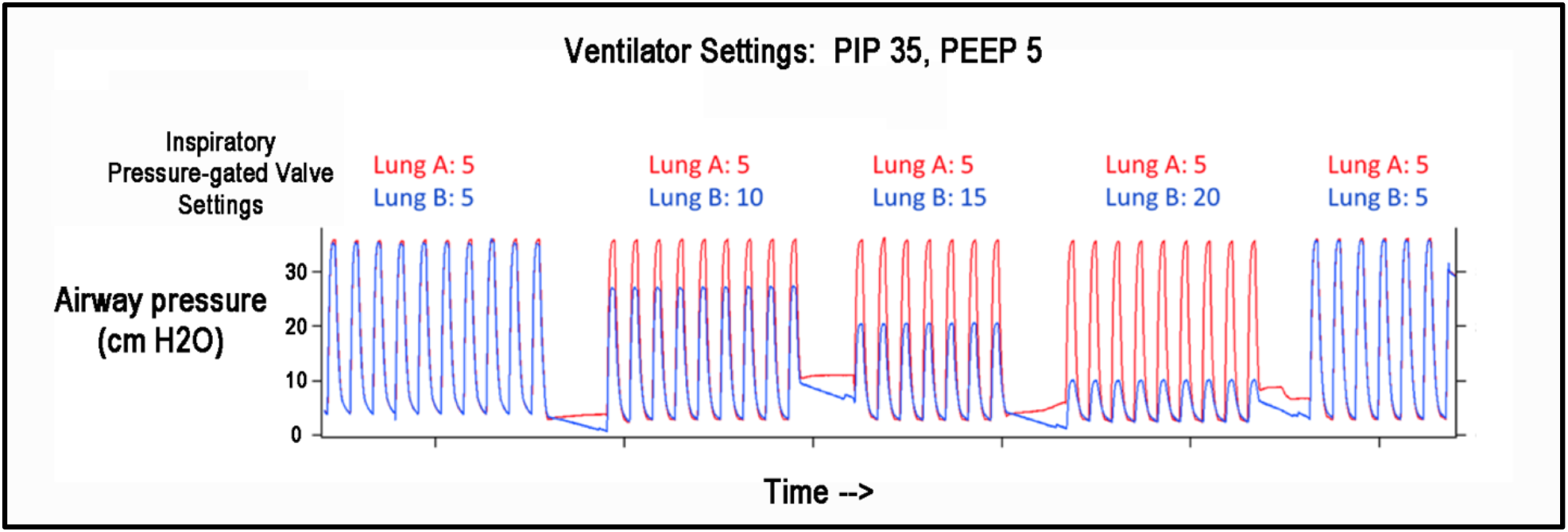
Impact of pressure-gated valve settings in inspiratory limb on ventilation pressures for two “patients”. Patient #1 corresponds to **Lung A**, while Patient #2 corresponds to **Lung B**. Inspiratory valve pressure settings are in cmH_2_O.

**Figure 11:**
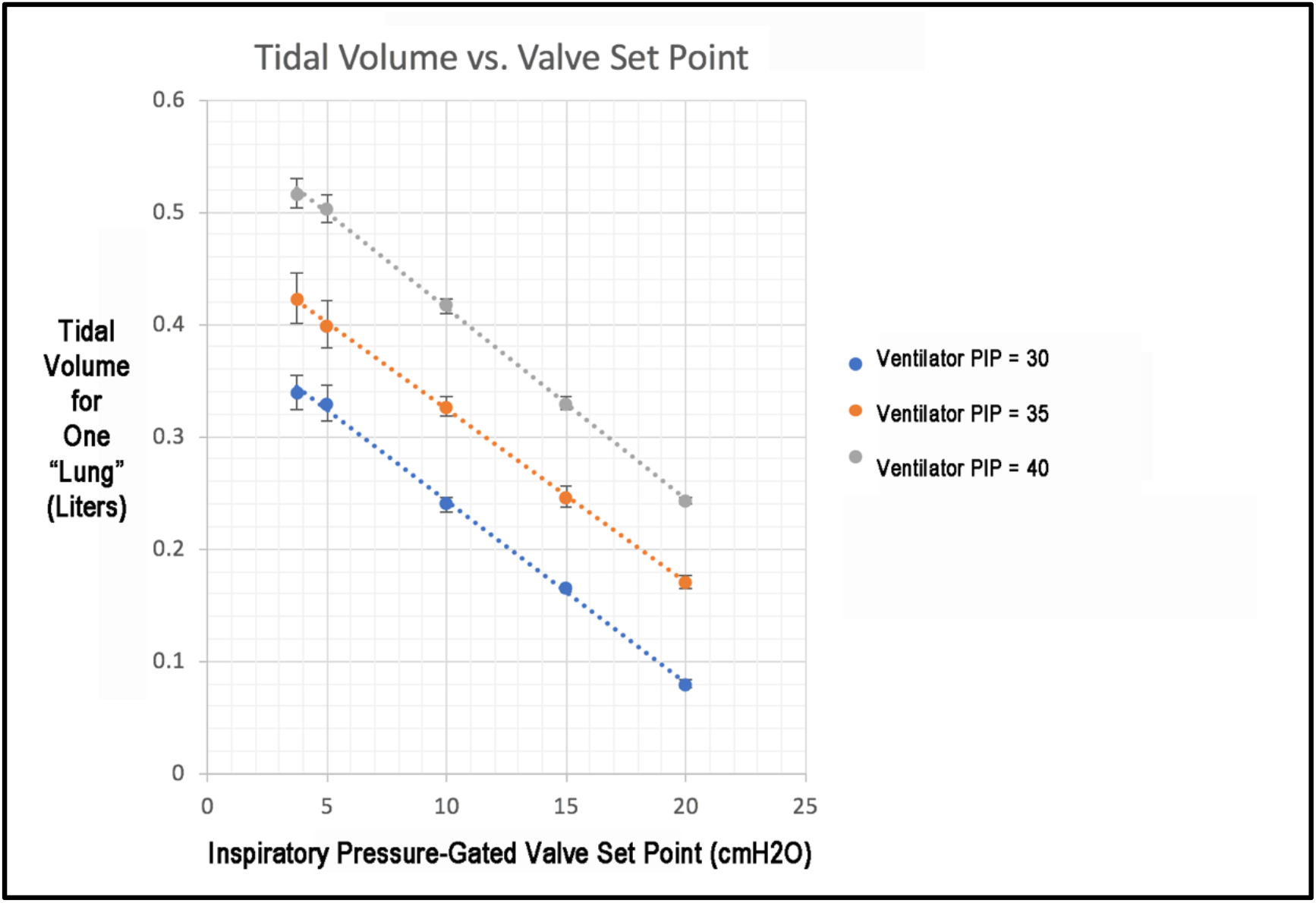
Impact of Inspiratory pressure-gated valve setting on tidal volume. Tidal volumes shown for 3 different settings of ventilator PIP. Data are from N=3 replicates. Dotted lines are linear fits.

**Figure 12:**
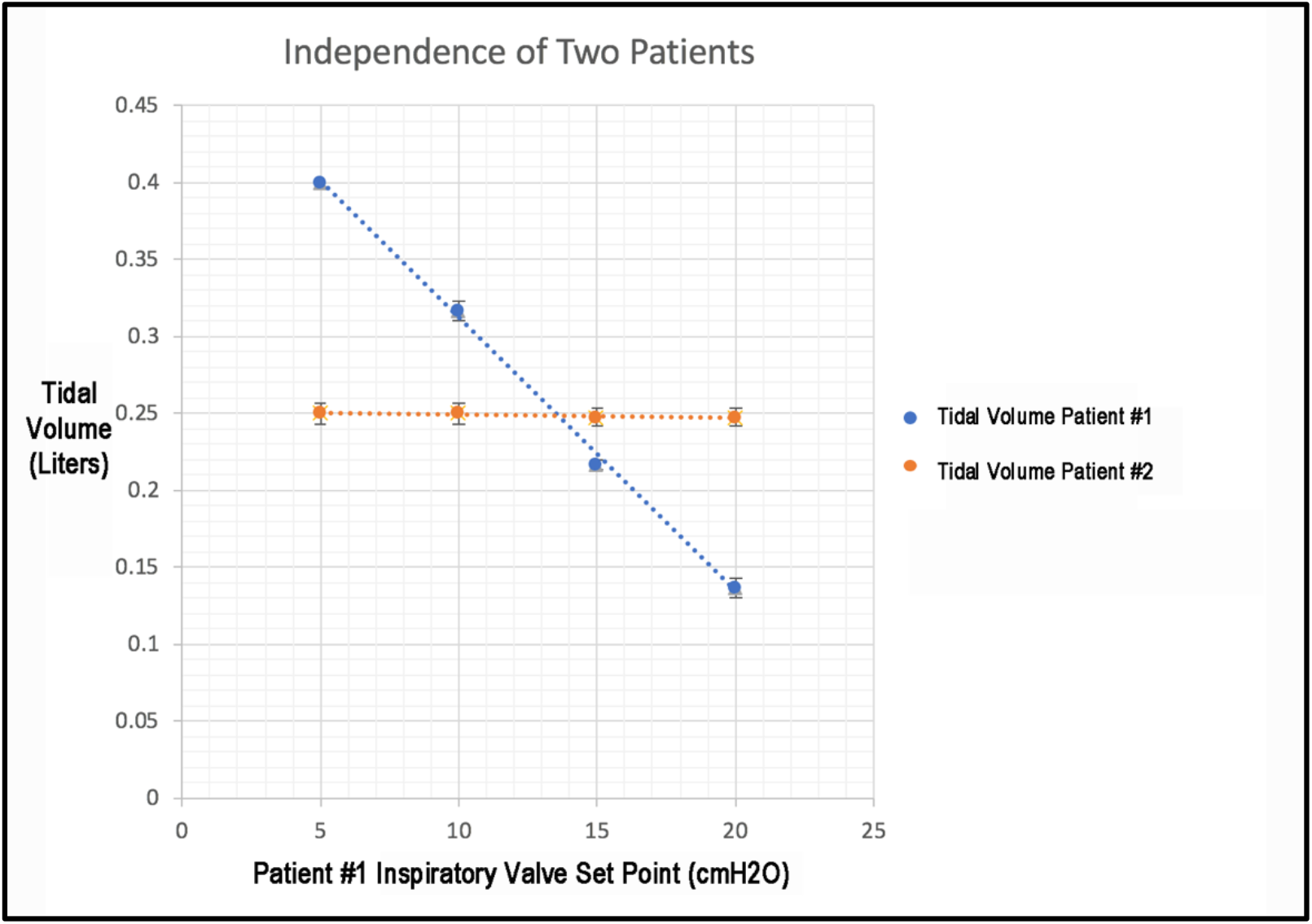
Impact of adjusting one inspiratory limb pressure-gated valve on the tidal volumes of Patients #1 and #2. Tidal volumes for both Patients shown for 4 different settings of the pressure-gated valve for Patient #1. Data are from N=3 replicates. Dotted lines are linear fits.

**Figure 13:**
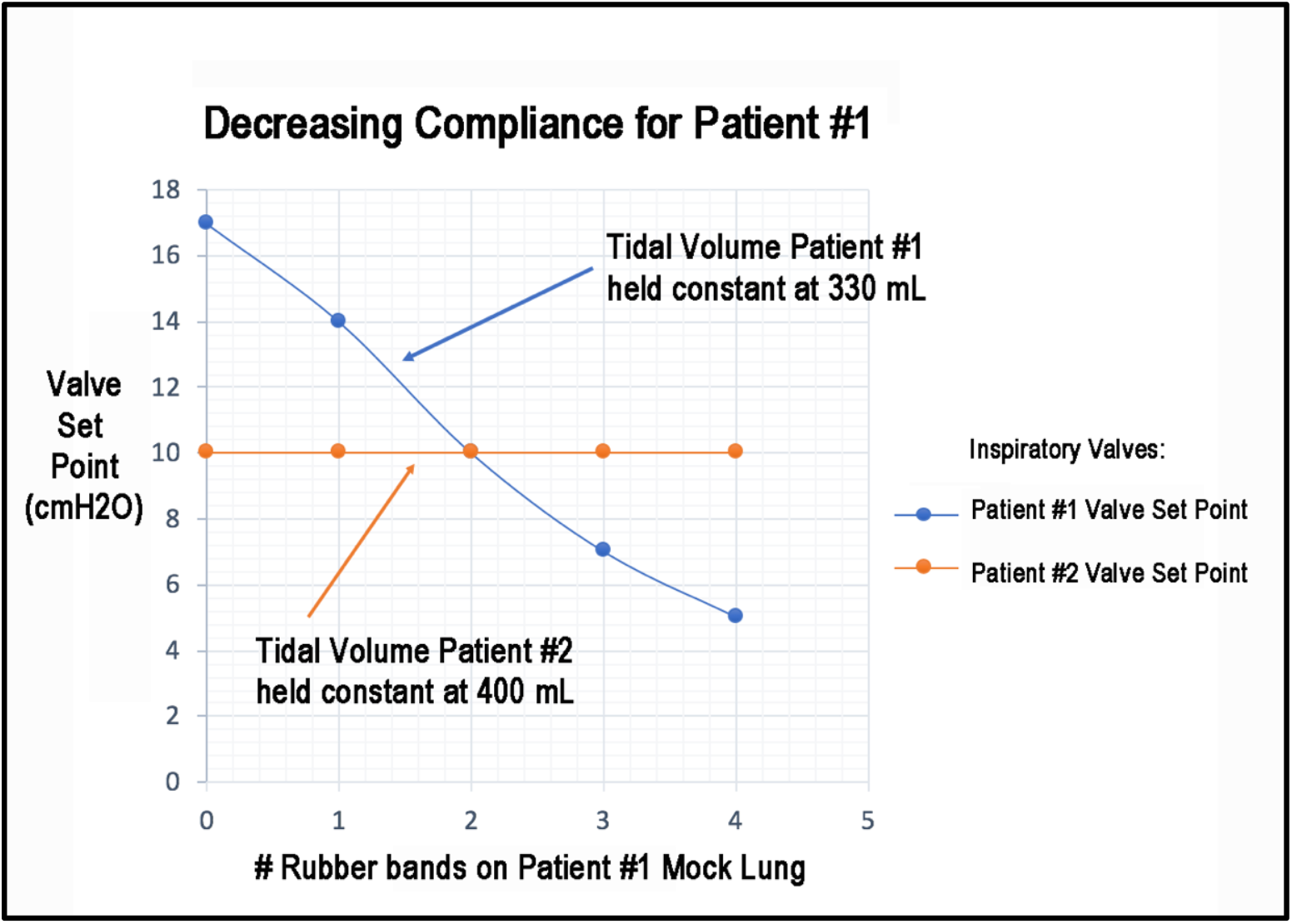
Impact of Decreasing Lung Compliance in One Patient. Compliance of mock lung for Patient #1 was decreased by sequentially adding rubber bands to the outside of the rubber ventilator bag. As Patient #1 compliance decreased, tidal volume could be maintained for Patient #1 by adjusting the pressure-gated valve for Patient #1 on the Inspiratory limb. During these maneuvers, tidal volumes for Patient #2 were not affected.

**Figure 14:**
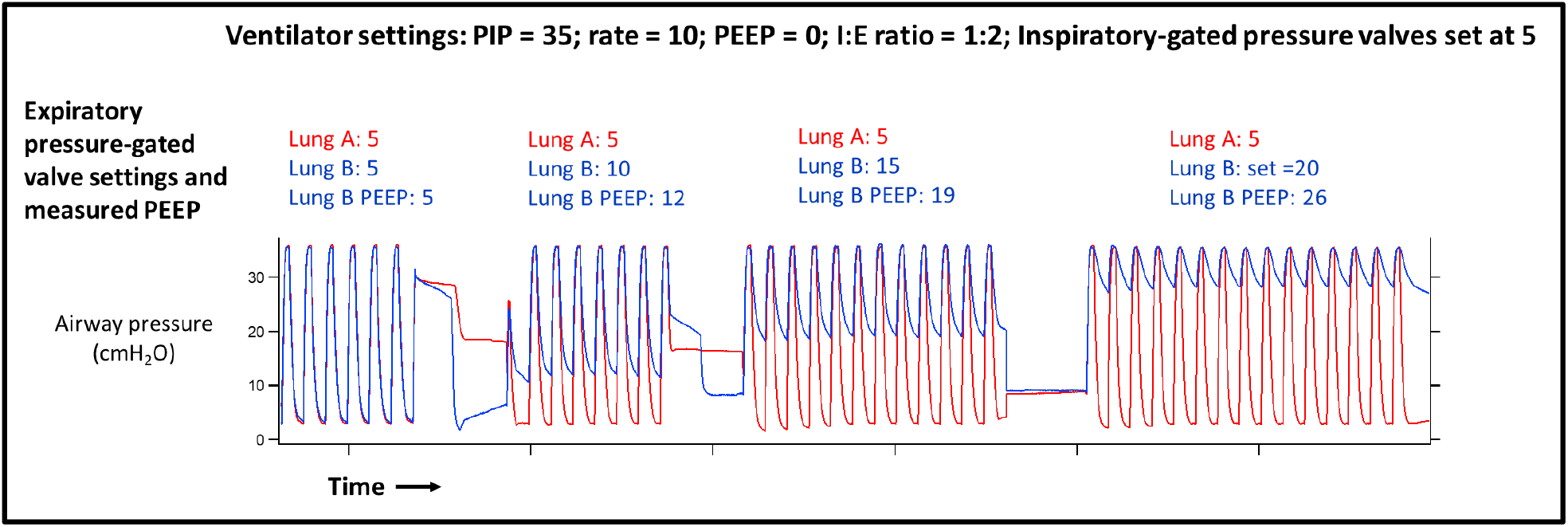
Impact of increasing the set point of the expiratory pressure-gated valve for one patient in the PReVentS circuit. By adjusting the set point of the expiratory pressure-gated valve for **Lung B**, we can selectively increase the amount of Positive End-Expiratory Pressure (PEEP) for **Lung B**, while not affecting **Lung A**. Data suggest the potential for “breath stacking” as indicated by higher levels of measured PEEP for **Lung B**. Patient #1 corresponds to **Lung A**, while Patient #2 corresponds to **Lung B**. Expiratory valve pressure settings are in cmH_2_O.

### I. Experiment #1: Determining Impact of Inspiratory Pressure-Gated Valves on Inspiratory Pressures

For this experiment, the circuit was connected to two mock lungs having similar compliance. The goal of the experiment was to tune the pressure settings in the pressure-gated valves in the inspiratory circuit, and to measure the resultant airway pressures for the two simulated “patients” (Figure 10). As described above, the inspiratory pressure for each patient is set by subtracting the setting of that patient’s inspiratory pressure-gated valve from the ventilator’s peak inspiratory pressure. Hence, for a set ventilator PIP of 35, a pressure-gated valve setting of 15 for Lung B results in an inspiratory pressure of 20 for that lung. Of note, the airway pressures of Lung A were entirely unchanged when varying the PIP of Lung B.

### II. Experiment #2: Determining Impact of Inspiratory Pressure-Gated Valves on Tidal Volumes

For this experiment, the circuit was connected to two mock lungs having similar compliance. The goal of the experiment was to tune the pressure settings in the pressure-gated valves in the inspiratory circuit, and to measure the resultant tidal volume for one simulated lung (Figure 11). As is clear from Figure 11, changing pressure settings on valves in the inspiratory limb of the PReVentS circuit provides a linear change in the resultant tidal volume to the lung, as would be expected for a simple rubber ventilator bag having relatively linear compliance.

As described above in Figure 9, setting the inspiratory pressure-gated valves to a given pressure results in a subtraction of that pressure from the ventilator inspiratory pressure for each patient. Hence, for a set ventilator PIP of 35 cmH_2_0, a pressure-gated valve setting of 15 cmH_2_0 for Lung B results in an inspiratory pressure of 20 cmH_2_0 for that lung. As the setting on the pressure-gated inspiratory valve increases, the resultant inspiratory pressure for that lung decreases, and the resultant tidal volume also decreases, in a linear fashion in this experiment.

### III. Experiment #3: Determining Impact of Inspiratory Pressure-Gated Valve Adjustment for One Patient Only

For this experiment, the circuit was connected to two mock lungs having similar compliance. The goal of the experiment was to tune settings of the inspiratory pressure-gated valve for one lung, and to measure the resultant tidal volumes for both of the lungs in the PReVentS circuit (Figure 12). As is clear from Figure 12, changing the pressure setting on one valve in the inspiratory limb for Patient #1 had a linear effect on the resultant tidal volume of Patient #1, while it had minimal to no effect on the measured tidal volume for Patient #2. The PReVentS circuit provides a linear change in the resultant tidal volume of Patient #1, as would be expected for a simple rubber ventilator bag having relatively linear compliance.

As described above in Figure 9, setting the inspiratory pressure-gated valves to a given pressure results in a subtraction of that pressure from the ventilator peak inspiratory pressure for each patient. As the setting on the pressure-gated inspiratory valve for Patient #1 increases, the resultant inspiratory pressure for Patient #1 decreases, and the resultant tidal volume for Patient #1 also decreases. Importantly, the tidal volume delivered to Patient #2 is unchanged by these maneuvers, since the ventilatory pressures to Patient #2 are not affected by these maneuvers.

### IV. Experiment #4: Determining Impact of Changing Compliance in One Patient

For this experiment, the circuit was connected to two mock lungs having similar compliance, and then compliance in one of the lungs was changed in increments, by applying rubber bands around the outside of the ventilator bag. The goal of the experiment was to determine the feasibility of maintaining tidal volumes in a patient with decreasing lung compliance, using the pressure-gated valves on the inspiratory limb for that patient to tune inspiratory pressure (Figure 13). As is clear from Figure 12, decreased compliance in Patient #1 could be largely offset, in terms of resulting tidal volume, by titrating the pressure on for Patient #1 on the inspiratory limb. In this study, the tidal volume for Patient #1 was maintained at approximately 330 mL, by means of decreasing the pressure-gated valve set point on the inspiratory limb from 17 to 5 cmH_2_O as the lung compliance fell. But despite the fact that the setting on the pressure-gated inspiratory valve for Patient #1 was being decreased, the inspiratory pressure and tidal volumes for Patient #2 remained unchanged. This experiment illustrates the potential feasibility of maintaining tidal volumes in one patient with changing lung mechanics, while not affecting the ventilation of the other patient treated with the PReVentS circuit.

### V. Experiment #5: Determining Impact of Changing Applied PEEP by Adjusting Pressure-Gated Valves on the Expiratory Limb

For this experiment, the circuit was connected to two mock lungs having similar compliance, and then the effective PEEP applied to Patient #2 (Lung B, Figure 14), was increased by increasing the set point of the pressure-gated valve on the expiratory limb for Patient #2. The goal of this experiment was to determine the impact of changing effective PEEP of one patient on ventilatory pressures and gas flows. As is clear from Figure 15, adjusting the set point of the expiratory valve for Patient #2 (“Lung B”) from 5 to 20 cmH_2_O did not affect the ventilation pressures for Patient #1 (“Lung A” in Figure 15). In contrast, the effective PEEP for Patient #2 was increased from 5 to approximately 25 cmH_2_O by increasing the set point on the expiratory valve for Patient #2 from 5 to 20 cmH_2_O.

As can be seen from careful inspection of Figure 14, there is some evidence of incomplete exhalation, or “breath stacking”, in the mock lung used to simulate Patient #2 in this experiment. Whether this observation is inherent to the PReVentS circuit, or whether it is an artifact of the specific circuit components or the mock lung utilized for this study, is unclear at the present time.

## Draft Protocol for Patient Management

**NOTE**: This draft protocol has not been tested on animals or patients. It is intended only as a starting point for thinking about how PReVentS could be valuable in the event that ventilator sharing became superior to all other options. Any consideration of ventilator sharing (using this circuit design or any other) must include a thorough evaluation of the technical, medical, legal, and ethical ramifications each time it is instituted.

### Important Information and Patient Safety Considerations

- Patients should be pharmacologically paralyzed and ventilator trigger settings should be maximized so that only mandatory breaths are delivered by the ventilator.
- Pressure-control ventilation must be used.
- All valves must be in the proper place, inserted in the proper orientation and functioning properly (neither sticking nor leaking).
- Certain ventilator alarms will *not* work as they do in single-patient operation, and it is critical to implement a comprehensive monitoring and alarm strategy and train all caretakers on it.
- Inspiratory pressures, expiratory pressures, and expiratory volumes should be measured independently for each patient at regular intervals. Ideally, these measurements should be continuous. Even judiciously set alarms on the shared ventilator will not be able to identify certain potentially serious changes in patient status.
- Valves in the patient circuit have the potential to limit flow even when open. Therefore, certain PReVentS circuits (depending on the specific ventilator, tubing, and valves used) might have reduced inspiratory flow or expiratory flow when compared to a traditional circuit. This would have the effect of slowing both inspiration and expiration, and could lead to reduced inspiratory volume or breath stacking at a given pressure setting and respiratory rate. While these occurrences can be adjusted for if they do occur, the provider must be vigilant for them and know how to respond appropriately.
- As the circuit is currently designed, negative circuit pressure near the patient will entrain room air through the negative-pressure relief valve – this will briefly *decrease the FiO2* following in-line suctioning, or if the patient makes strong respiratory effort during the ventilator’s expiratory phase. If even a brief FiO2 decrease is unacceptable for a patient, an oxygen reservoir bag should be added at the negative pressure relief valve. With proper forethought, steps can also be taken to mitigate FiO2 drop during in-line suctioning (see below).
- While not intrinsic to the PReVentS circuit, with the current hand-made valves it is necessary to briefly open the circuit to make valve adjustments. During this maneuver, ventilation is still maintained for the other patient, and PEEP is maintained for both patients. We are currently iterating on valve design to allow pressure adjustment without any circuit disruption.

#### I. Monitoring

When sharing a single ventilator between two patients, the inbuilt safety monitors of the ventilator will not function as they usually do. It is therefore necessary to develop and implement a monitoring strategy that will reliably alert providers to unexpected changes and dangerous events. In the current crisis, not just ventilators but many types of medical devices are in short supply, which may affect the ability to provide monitoring. With that in mind, we propose a minimal monitor setup (that is absolutely required and will identify all the changes in patient condition we have so far considered) and a recommended monitor setup (which provides redundancy, could identify unconsidered situations, and would make patient management significantly easier and safer). The decision to proceed with only limited monitoring would necessarily depend upon the available alternatives and perceived balance of risk and benefit. This monitoring strategy is indebted to those already published by other groups considering ventilator splitting (Beitler 2020, Pinson 2020).

##### Minimal Monitors

1. Shared ventilator – Measures and displays sum of patient tidal volumes (VTe), summed minute ventilation (Ve), and volume delivered by ventilator with each breath (Vdel). Additionally, it can identify certain disconnects and occlusions. We believe that a carefully set VTe alarm can identify most situations that require provider attention with good sensitivity and specificity (see below and Appendix S1). Furthermore, in the absence of superior alternatives, individual patient tidal volumes can be measured with the shared ventilator by briefly occluding the other patient’s circuit (which puts the occluded patient into an expiratory hold). If measuring independent patient VTe in this manner, be sure to note that many ventilators adjust this measurement for the circuit compliance. Therefore, tidal volume may be mis-estimated by 80ml (Beitler 2020). The addition of the bias flow short circuit may also contribute to the shared VTe, but this can be minimized with a small and non-compliant bypass. This bypass volume can also be accounted for by clamping each patient one at a time and solving for the bypass circuit VTe algebraically.
2. Individual manometers near each patient Y-piece to monitor individual airway pressures – These measure peak inspiratory pressure (PIP) and positive end-expiratory pressure (PEEP) for each individual patient. Ideally, these manometers should provide information continuously and be alarmable (see below). At minimum, it is important to spot check airway pressures every time ventilator parameters are changed and whenever the Vte alarm identifies a change in system status.
3. End-tidal CO_2_ measurement – To ensure appropriate ventilation, patient CO_2_ should be monitored with arterial blood gas analysis, capnography, or a combination. This type of monitoring is typically standard for each ICU patient.
4. Standard ICU monitors (pulse oximetry, blood pressure, EKG, temperature).

##### Recommended Monitors

1. Continuous manometry at each patient Y-piece: This may be most easily done by attaching a standard invasive blood-pressure transducer to a connector placed near the patient Y-piece (Pinson 2020). Airway pressures can then be displayed on a patient’s vital signs monitor, appropriate alarms can be set to assure appropriate PIP and PEEP, and this information may even be remotely available at the nurses’ station or elsewhere on the hospital floor. Note that pressures measured this way may be reported in mmHg rather than cmH_2_O, so conversion tables and provider training should be provided.
2. Independent tidal volume monitoring: This could be accomplished by a freestanding respiratory monitor with the flow sensor just distal to the patient Y-piece (Beitler 2020).However, these appear to be a relatively limited resource. We are actively in the process seeing whether outpatient pneumotachometers can be adapted to this purpose. Another alternative is that some ICU ventilators allow for attachment of an external flow sensor (for instance, Carefusion markets the VarFlex sensor) which could be used to measure a single patient’s VTe, and the other patient’s could be approximated by subtracting the first patient’s VTe from the combined VTe from the shared ventilator.
3. Continuous capnography of each patient: could identify accidental hypo- or hyperventilation in real time and alert the provider.

##### Proposed Alarm Settings

· Shared VTe: Low alarm = Shared VTe – 50 ml, High alarm = Shared VTe + 50 ml, alarm set to trigger if 2 breaths are outside of this range
· Individual VTe (if available): Low alarm = VTe – 50 ml, High alarm = VTe + 50 ml
· Ventilator Ve: Low alarm = Ve – 1 L, High alarm = Ve + 1 L
· Ventilator RR: Low alarm = RR – 3 bpm, High alarm = RR + 3 bpm
· PIP alarm (if available): Low alarm = Set PIP – 5 cmH_2_O, High alarm = Set PIP + 5 cmH_2_O
· PEEP alarm (if available): Low alarm = Set PEEP – 2 cmH_2_O, High alarm = Set PIP + 5 cmH_2_O
· ETCO_2_ alarm (if available): Low alarm = Target ETCO_2_ – 5 mmHg, High alarm = Target ETCO_2_ + 5 mmHg

#### II. Patient Care

Because ventilation with the PReVentS paradigm relies upon setting individual patient inspiratory pressures and expiratory pressures (similar to any pressure-controlled ventilation mode on a single patient ventilator), most adjustments should be familiar to the provider. However, the described circuit and the necessary patient monitoring departs enough from normal practice that it is worthwhile to describe how we propose certain common tasks might be performed.

##### Adjusting PIP or Tidal Volume

Assuming the ventilator PIP (Inspiratory Pressure + PEEP) is set at 40 cmH_2_O, each patient’s PIP is set independently by the inspiratory limb pressure gated valve, and it will be equal to 40 minus the valve setting. To adjust this PIP for one patient, make the desired adjustment on the inspiratory limb pressure gated valve (decrease the valve setting to increase PIP and VT, increase valve setting to decrease PIP and VT). Measure this patient’s airway pressures to make sure they reflect the desired change. Measure this patient’s VTe to make sure it is appropriate. View flow tracings on the ventilator or pressure tracings on the patient monitor to alert you to breath stacking. Adjust alarms to reflect the new settings.

##### Adjusting PEEP

Assuming the ventilator PEEP is set at 0 cmH_2_O, each patient’s PEEP is set independently by the expiratory limb pressure gated valve. To adjust this PEEP, make the desired adjustment on the expiratory limb pressure gated valve (decrease valve setting to decrease PEEP, increase valve setting to increase PEEP). After this change, consider if you need to adjust the same patient’s PIP to maintain a constant driving pressure or tidal volume. Measure this patient’s airway pressures to make sure they reflect the desired change. Measure this patient’s VTe to make sure it is appropriate. View flow tracings on the ventilator or pressure tracings on the patient monitor to alert you to breath stacking. Adjust alarms to reflect the new settings.

##### Other Ventilation Changes

Other ventilation parameters, including RR and FiO_2_, are shared between all patients sharing the ventilator. Beitler and colleagues provided sound recommendations for tailoring these ventilator settings to the care of multiple patients on a single ventilator (Beitler 2020). Even though respiratory rate is shared, the PReVentS paradigm allows for differential minute ventilation by individualizing patient tidal volumes. Whenever respiratory rate is adjusted, airway pressures and tidal volumes should be confirmed, alarms should be adjusted, and CO_2_ should be measured at an appropriate interval.

##### Removing a Patient from the Circuit

A patient can be removed from the circuit with minimal effect on the other patient. To do so, clamp the inspiratory limb of the circuit leading to the patient to be removed. The ventilator will continue to ventilate the other patient normally. The patient can then be removed from the circuit either at the ETT, or (to maintain recruitment) along with the piece of circuit containing the inspiratory check valve and expiratory pressure-gated valve. Adjust the alarm parameters on the ventilator to reflect that only one patient is being ventilated.

##### ETT Suctioning

If suctioning a patient with an in-line suction catheter, negative pressure generated near the patient Y-piece will open the negative pressure relief valve and entrain room air (see discussion above). If suction is sufficiently strong, it may even reduce the inspiratory pressures of the other patient during suctioning. To minimize the risk of hypoxia, increase FiO_2_ prior to suctioning. If necessary, you can also attach an oxygen reservoir to the negative pressure relief valve.

#### III. Anticipated Events, Effects, and Alarms

It is important to address various conceivable deviations, what effects these events would have on the patients attached to the ventilator, and how the provider would be alerted to these occurrences. We have attempted to assemble a comprehensive list of such events and their effects (Appendix S1). A brief summary of some anticipated events and failure modes is presented here.

##### Decreased Lung Compliance

A variety of clinical changes could lead to decreased compliance – mucous plug, kinked ETT, breath stacking, pneumothorax, hemothorax. While these events can pose a profound risk to the affected patient, they will have no effect on the respiration of other patients sharing a PReVentS circuit. Decreased compliance in one patient circuit would reliably decrease the VTe measured at the ventilator, and would trigger an alarm.

##### Increased Lung Compliance

The most likely cause of increased compliance in the acutely ill would be lung recovery, or perhaps collection drainage, though other causes such as sudden onset of flail chest could occur. None of these events would change the ventilation of patients sharing the ventilator. Increased compliance would increase VTe and trigger an alarm.

##### Circuit Leak/Disconnect

The effect of a circuit leak on the PReVentS circuit will depend upon where the leak occurs (Appendix S1). In general, if the leak is within a single patient circuit (anywhere between the inspiratory pressure gated valve and the expiratory check valve), other patients sharing the ventilator will maintain their PEEP and receive some degree of ventilation. The closer the leak is to the expiratory pressure-gated valve, the more normally other patients will be ventilated. All leaks from the circuit will trigger the low VTe alarm, and some will trigger the circuit disconnect alarm as well.

Of note, if almost any valve within the circuit develops a leak, it will trigger the high VTe alarm. The one exception is the negative pressure relief valve, which will act as a circuit leak and trigger the low VTe alarm.

##### Circuit Occlusion/Stuck Valve

The effects of a circuit occlusion are also dependent on the occlusion location (again, see Appendix S1). However, any occlusions on a patient circuit between the inspiratory T-piece and the expiratory T-piece will have no effect on the ventilation of other patients sharing the ventilator. All circuit occlusions will trigger the low VTe alarm, and some will trigger an occlusion alarm as well.

Most stuck valves will register as circuit occlusions, with the exception of negative pressure relief valves. An occlusion at this valve could go unnoticed until a patient is suctioned or makes a respiratory effort. In that case, the affected patient might experience negative airway pressures until the next inspiratory phase. It is therefore necessary that the negative pressure relief valve in each patient circuit is tested before initiating ventilation with a PReVentS circuit.

##### Patient Ventilatory Effort

While it is important that patients not spontaneously breathe for the PReVentS protocol, it is also important that the system be robust to unanticipated ventilatory effort. Depending upon how ventilatory efforts align with the ventilator cycle, the effects on the spontaneously breathing patient will be variable (Appendix S1). In general, the spontaneously-breathing patient’s tidal volumes will increase, and room air may be entrained through the negative pressure relief valve into that patient’s circuit. We anticipate that there will be no effect on the other patient’s ventilation or FiO2. Respiratory effort will trigger the high VTe alarm.

##### Attempted Chest Compressions

We do not recommend that chest compressions be performed upon a patient attached to a shared ventilator – with our proposed circuit design, it would be easy and safe to disconnect the patient in arrest without releasing an infected aerosol or interrupting ventilation to the paired patient. However, it is possible that one patient may undergo chest compressions while the patient is still attached to the ventilator. In our laboratory testing, simulated chest compressions on one patient attached to the PReVentS circuit led to only minor changes in airway pressures for the other patient, and the ventilatory pattern to the second patient remained regular.

## Conclusions

The PReVentS circuit and protocol may provide a means to care for multiple patients, all having severe lung disease, with one ventilator. Unlike previously described approaches, the tailoring of ventilatory pressures for each patient, and the ability to titrate those pressures over time, may provide a more useful means of stretching ventilator resources.

The PReVentS circuit design must still be tested in animals and in patients. Such testing will almost certainly bring about additional system improvements and modifications. We fully expect to improve the means of set point adjustment for in-line valves in the circuit. Furthermore, additional in vivo experimentation will clarify the best monitors and strategies to assure the safety of both patients attached to the ventilator. As such, we anticipate that ongoing improvements and reports on this system will follow this initial report.

## Data Availability

All data will be provided upon reasonable request.

## Acknowledgements

The authors want to acknowledge extremely helpful conversations with Dr. Roberta Hines and Dr. Naftali Kaminski. Additionally, we would like to acknowledge the help and support of the respiratory therapy team at Yale-New Haven Hospital.

